# PHYSICAL ACTIVITY AND ASSOCIATED FACTORS AMONG PATIENTS ATTENDING HYPERTENSION CLINICS IN MBARARA CITY, SOUTHWESTERN UGANDA

**DOI:** 10.1101/2023.06.28.23292015

**Authors:** Atwongyeire Night, Ssewanyana Anna Maria, Namukwaya Racheal, Mutesasira Edward, JJunju Fred, Nuwahereza Amon, Niyonsenga Jean Damascene, Zillah Whitehouse, Kazibwe Herman, Arubaku Wilfred

## Abstract

**Background:** Physical activity has been shown to prevent mortality and morbidity among people with hypertension. Hypertension has been identified to affect about 33% of the adult population across the globe and 26.4% in Uganda. An increase in hypertension and its adverse outcomes have been observed and reported in Southwestern Uganda. This may be due to physical inactivity. However, there is limited evidence demonstrating physical activity among patients with hypertension within Southwestern Uganda. The current study investigated the extent of physical activity and associated factors among patients attending hypertension clinics in Mbarara City, Southwestern Uganda.

**Objective:** To investigate the extent of physical activity and associated factors among patients attending hypertension clinics in Mbarara City, Southwestern Uganda.

**Methods:** A descriptive, quantitative cross-sectional study was conducted. Participants’ data was obtained using a structured researcher-administered questionnaire consisting of the International Physical Activity Questionnaire (IPAQ) long form, Motivation for Physical Activity Questionnaire (RM4-FM) and the Barrier to Being Active Quiz (BBAQ). Frequency distribution tables, Fischer’s exact test and multivariate logistic regression were used to describe data and establish associations during data analysis. A p-value less than 0.05 with 95% confidence interval was considered to be statistically significant.

**Results:** Less than half of the participants (45.39%) were found to be physically active. Level of education, having heard of physical activity, place of residence, sedentary behaviour and social influence were associated with physical activity with the Fischer’s exact test (p-value<0.05). On multivariate adjustment, only level of education (aOR=1.374; CI=1.055-1.790; p-value=0.018) and sedentary behaviour (aOR=0.276; CI=0.126-0.606; p-value=0.001) remained significant factors associated with physical activity. Majority of those who were active reported to be autonomously motivated. Most reported barriers were lack of skill, social influence and lack of willpower.

**Conclusion:** More than half of the patients with hypertension were physically inactive and almost half were inactive and sedentary. This is a double disaster especially for patients with hypertension; therefore, emphasis should be put on educating the public about the benefits of physical activity and availing a variety of physical activity options that engage all demographic groups.

## Introduction

Across the globe, 7.6% of deaths due to cardiovascular diseases are attributed to physical inactivity (1). Evidence also shows that 74.0% of the cardiovascular disease deaths associated with physical inactivity occurs in low and middle-income countries. Physical activity has been recommended in management of hypertension (1). In 1968, one of the first clinical studies demonstrating the antihypertensive effect of physical activity was published. The study found a 13.4mmHg decrease in the mean systolic pressure and a 2 to 30 mmHg decrease in diastolic pressure, after the completion of the exercise training period (2). More studies have recently been done to confirm the effect of physical activity on blood pressure control through various mechanisms (3) including, decreasing sympathetic activity (4) and reducing oxidative stress (5). The notion that an individual can be both physically active and also sedentary (6) has come as an eye-opener for most physical activity researchers, and some findings have examined the possible negative effect of sedentary behaviour over physical activity in physically active populations (7) and sedentary behaviour has been reported to produce direct negative effects on outcomes of hypertension (8). Evidence comes from (9) who reported that each one hour of sedentary behaviour was associated with 0.06mmHg increase in systolic pressure and 0.20mmHg increase in diastolic pressure among hypertensives. In addition, sedentary behaviour has been reported to compromise outcomes of hypertension leading to stroke and myocardial infarction among others (10). Despite the known benefits of physical activity, physical inactivity especially among individuals with hypertension is still high across the globe ranging from 23% in adults to 81% in adolescents (11). It is important to note that the prevalence of physical inactivity varies across various regions and countries and may be as high as 80% among some adult populations (12). The major form of motivation to physical activity adherence has been reported to be autonomous motivation (13) Additionally, (14) found that autonomous regulation was the main predictor of exercise intensity, frequency and time among males and females. The major barriers that have been reported include inadequate information about physical activity, fear of physical activity, health illiteracy and fear of changing lifestyle (15).

In East Africa, in a study in Kenya, it was found that majority (63.0%) of the patients with hypertension had low levels of physical activity (16). In another study done in Kigali Rwanda, it was indicated that the major factors associated with physical activity among patients with hypertension were age, marital status, place of residence, and education level, among others, and the perceived motivators were perceived health benefits, self-efficacy and blood pressure control (17)

On that account, increasing incidences of adverse outcomes of hypertension like heart failure, renal failure and stroke among individuals with hypertension have been reported and observed in this region (18-20). It is important to note that, in a recent study by (21) at Mbarara Regional Referral hospital, hypertension was cited as a major contributing factor to stroke mortality. These observations could be attributed to poor practice of lifestyle modifications such as physical inactivity among patients with hypertension. However, this argument cannot be fully supported since there is still limited evidence on physical activity among individuals with hypertension in this region. Relatedly, there is inadequate information on the factors associated with physical activity that might influence the extent of physical activity in this population. Therefore, this study sought to investigate the extent of physical activity and associated factors among patients attending hypertension clinics in Mbarara City, Southwestern Uganda.

## Methods

### Study design

A descriptive, quantitative cross-sectional study using a researcher-administered questionnaire was conducted. Hypertensive patients were identified by the nurse and connected to the research members, and a consecutive sampling strategy was used to recruit participants in the study. Interviewer-administered questionnaires were adopted from the International Physical Activity questionnaire (IPAQ) long form to investigate the extent of physical activity, Motivation for physical activity (RM4-FM) to explore motivators to physical activity and the Barriers to Being Active Quiz (BBAQ) to explore barriers to physical activity.

### Study setting

The study was conducted at two hypertension clinics in Mbarara City, Southwestern Uganda. The two clinics were DMA Diagnostics and Laboratory Limited in Kamukuzi division and Mbarara Medical Specialist clinic located in Kakoba division. The participants got from these clinics were 133 patients and 99 respectively, obtained through proportionate sampling.

### Study variables

The level of physical activity (dependent variable) was analyzed by considering the metabolic equivalent of the task, METs-minutes per week and the days of physical activity. MET-minutes per participant were calculated using the formula (MET value) * (time of activity in minutes per day* number of days the activity was done in a week). The independent variables were the associated factors to physical activity. These included: socio-demographic characteristics (including age, gender, place of residence, monthly income, occupation, marital status, level of education), duration of hypertension, awareness of physical activity, source of information about physical activity, sedentary behaviour, motivators for physical activity, barriers to physical activity and sedentary behaviour. Sedentary behaviour was measured from time spent sitting as average sitting time in the past seven days, assessed by the International Physical Activity Questionnaire (IPAQ). Motivators and barriers were measured using Motivation for Physical Activity Questionnaire (RM4-FM) and Barriers to Being Active Quiz (BBAQ)

### Data analysis

Given that the total study sample size was small, Fischer’s exact test was used to test for association between physical activity and the independent variables. Multivariate logistic regression was also used to test for further association between physical activity and the independent variables. A p-value less than 0.05 was considered for statistical significance.

## Results

Overall, 141 participants participated in the study. Median age of the participants was 58 years (IQR=14). *Table 1* shows that females accounted for 67.4% of the participants while males accounted for 32.6% and the age categories were: less than 35 years 3.6%, 35-49 years 18.4% and 50 years above 78.0%. The places of residence of the participants were: Mbarara 44.0%, Greater Bushenyi 15.6%, Isingiro 13.5% and others 27.0%. The different occupations for the participants were: Farming 27.0%, business 23.4%, housewife 10.6%, retired 9.2% and others 29.8%. The monthly income of the participants was 0-500,000UGX 55.3%, 500,000-1M UGX 31.9% 1M UGX or more 12.8% while the marital status of the participants was: Married 73.8% single 6.4% divorced 5.7% and others 14.2%. The duration of hypertension among the participants was: less than one year 14.2%, 1-5 years 38.3% and more than 6 years 47.5%. The level of education of participants was; completed primary 22.0% secondary 19.9%, tertiary 34.0% and no formal education 24.1%. Those who had heard of physical activity were 85.8% and those who never heard of physical activity were 14.2%. Source of information (from whom) included friends and family 17.7%, health workers 56.0%, mass media 12.1%. The overall physical activity referred to a combination of activities in the different domains. *Table 2* shows that overall, of the 141 participants, 54.6% participants had low levels of physical activity, 40.4% achieved moderate levels of physical activity and 5.0% had high levels of physical activity. The factors found to be associated with physical activity on Fishers exact test included place of residence, social influence, level of education, information about physical activity and sedentary behaviour, as shown in *table 3*. On bivariate and multivariate logistic regression, only two factors remained statistically significant and these were; sedentary behavior and level of education, as shown in *table 4*.

**Table 1:** Demographic characteristics of study participants

| Characteristics | Frequencies | % |
| --- | --- | --- |
| **Age group** |  |  |
| less than 35 years | 5 | 3.6 |
| 35-49 years | 26 | 18.4 |
| 50 years and above | 110 | 78.0 |
| **Gender** |  |  |
| Female | 95 | 67.4 |
| Male | 46 | 32.6 |
| **Place of residence** |  |  |
| Greater Bushenyi | 22 | 15.6 |
| Isingiro | 19 | 13.5 |
| Mbarara | 62 | 44.0 |
| Others | 38 | 27.0 |
| **Occupation** |  |  |
| Business | 33 | 23.4 |
| Farmer | 38 | 27.0 |
| Housewife | 15 | 10.6 |
| Others | 42 | 29.8 |
| Retired | 13 | 9.2 |
| **Monthly income** |  |  |
| 0-500000 UGX | 78 | 55.3 |
| 500000-1M UGX | 45 | 31.9 |
| 1MUGX or more | 18 | 12.8 |
| **Marital status** |  |  |
| Divorced | 8 | 5.7 |
| Married | 104 | 73.8 |
| Other | 20 | 14.2 |
| Single | 9 | 6.4 |
| **Duration of hypertension** |  |  |
| 1-5 years | 54 | 38.3 |
| More than 6 years | 67 | 47.5 |
| less than one year | 20 | 14.2 |
| **Level of education** |  |  |
| No formal education | 34 | 24.1 |
| Completed Primary | 31 | 22.0 |
| Completed Secondary | 28 | 19.9 |
| Completed Tertiary | 48 | 34.0 |
| **Heard of physical activity** |  |  |
| No | 20 | 14.2 |
| Yes | 121 | 85.8 |
| **From whom** |  |  |
| Friends and family | 25 | 17.7 |
| Health worker | 79 | 56.0 |
| Mass media | 17 | 12.1 |
| Never heard of physical activity | 20 | 14.2 |
| **Total** |  |  |
| All | 141 | 100 |

**Table 2:** Extent of physical activity among the participants.

| Domains of physical activity | Frequencies | % |
| --- | --- | --- |
| **Job-related physical activity** |  |  |
| Low | 130 | 92.2 |
| Moderate | 10 | 7.1 |
| High | 1 | 0.7 |
| **Transportation physical activity** |  |  |
| Low | 131 | 92.9 |
| Moderate | 10 | 7.1 |
| High | 0 | 0.0 |
| **Housework and house maintenance** |  |  |
| Low | 124 | 87.9 |
| Moderate | 16 | 11.4 |
| High | 1 | 0.7 |
| **Recreation,sport and leisure-time physical activity** |  |  |
| Low | 125 | 88.7 |
| Moderate | 16 | 11.4 |
| High | 0 | 0.0 |
| **Overall physical activity score** |  | % |
| Low | 77 | 54.6 |
| Moderate | 57 | 40.4 |
| High | 7 | 5.0 |
| **Total** |  |  |
| All | 141 | 100 |

**Table 3:** Extent of physical activity and sociodemographic characteristics.

|  | Frequencies |  |  | Total | % |  |  | Total |  |
| --- | --- | --- | --- | --- | --- | --- | --- | --- | --- |
| Characteristic | Physical activity score |  |  | All | Physical activity score |  |  | All | p-value |
|  | Low | Moderate | High |  | Low | Moderate | High |  |  |
| **Age group** |  |  |  |  |  |  |  |  |  |
| less than 35 years | 4 | 1 | 0 | 5 | 80.0 | 20.0 | 0.0 | 100 | 0.549 |
| 35-49 years | 16 | 8 | 2 | 26 | 61.5 | 30.8 | 7.7 | 100 |  |
| 50 years and above | 57 | 48 | 5 | 110 | 51.8 | 43.6 | 4.5 | 100 |  |
| **Gender** |  |  |  |  |  |  |  |  |  |
| Female | 50 | 42 | 3 | 95 | 52.6 | 44.2 | 3.2 | 100 | 0.211 |
| Male | 27 | 15 | 4 | 46 | 58.7 | 32.6 | 8.7 | 100 |  |
| **Place of residence** |  |  |  |  |  |  |  |  |  |
| Greater Bushenyi | 7 | 15 | 0 | 22 | 31.8 | 68.2 | 0.0 | 100 | 0.016 |
| Isingiro | 15 | 3 | 1 | 19 | 78.9 | 15.8 | 5.3 | 100 |  |
| Mbarara | 31 | 26 | 5 | 62 | 50.0 | 41.9 | 8.1 | 100 |  |
| Others | 24 | 13 | 1 | 38 | 63.2 | 34.2 | 2.6 | 100 |  |
| **Occupation** |  |  |  |  |  |  |  |  |  |
| Business | 18 | 12 | 3 | 33 | 54.5 | 36.4 | 9.1 | 100 | 0.705 |
| Farmer | 20 | 17 | 1 | 38 | 52.6 | 44.7 | 2.6 | 100 |  |
| Housewife | 9 | 6 | 0 | 15 | 60.0 | 40.0 | 0.0 | 100 |  |
| Others | 25 | 14 | 3 | 42 | 59.5 | 33.3 | 7.1 | 100 |  |
| Retired | 5 | 8 | 0 | 13 | 38.5 | 61.5 | 0.0 | 100 |  |
| **Monthly income** |  |  |  |  |  |  |  |  |  |
| 0-500000 UGX | 46 | 31 | 1 | 78 | 59.0 | 39.7 | 1.3 | 100 | 0.166 |
| 500000-1M UGX | 22 | 19 | 4 | 45 | 48.9 | 42.2 | 8.9 | 100 |  |
| 1M UGX or more | 9 | 7 | 2 | 18 | 50.0 | 38.9 | 11.1 | 100 |  |
| **Marital status** |  |  |  |  |  |  |  |  |  |
| Divorced | 2 | 6 | 0 | 8 | 25.0 | 75.0 | 0.0 | 100 | 0.376 |
| Married | 58 | 39 | 7 | 104 | 55.8 | 37.5 | 6.7 | 100 |  |
| Single | 4 | 5 | 0 | 9 | 44.4 | 55.6 | 0.0 | 100 |  |
| Other | 13 | 7 | 0 | 20 | 65.0 | 35.0 | 0.0 | 100 |  |
| **Duration of hypertension** |  |  |  |  |  |  |  |  |  |
| less than one year | 11 | 8 | 1 | 20 | 55.0 | 40.0 | 5.0 | 100 | 0.825 |
| 1-5 years | 30 | 20 | 4 | 54 | 55.6 | 37.0 | 7.4 | 100 |  |
| More than 6 years | 36 | 29 | 2 | 67 | 53.7 | 43.3 | 3.0 | 100 |  |
| **Level of education** |  |  |  |  |  |  |  |  |  |
| No formal education | 24 | 10 | 0 | 34 | 70.6 | 29.4 | 0.0 | 100 | 0.026 |
| Completed Primary | 21 | 10 | 0 | 31 | 67.7 | 32.3 | 0.0 | 100 |  |
| Completed Secondary | 12 | 14 | 2 | 28 | 42.9 | 50.0 | 7.1 | 100 |  |
| Completed Tertiary | 20 | 23 | 5 | 48 | 41.7 | 47.9 | 10.4 | 100 |  |
| **Heard of physical activity** |  |  |  |  |  |  |  |  |  |
| No | 16 | 3 | 1 | 20 | 80.0 | 15.0 | 5.0 | 100 | 0.027 |
| Yes | 61 | 54 | 6 | 121 | 50.4 | 44.6 | 5.0 | 100 |  |
| **From whom** |  |  |  |  |  |  |  |  |  |
| Friends and family | 11 | 14 | 0 | 25 | 44.0 | 56.0 | 0.0 | 25 | 0.390 |
| Health worker | 42 | 31 | 6 | 79 | 53.2 | 39.2 | 7.6 | 79 |  |
| Mass media | 8 | 9 | 0 | 17 | 47.1 | 52.9 | 0.0 | 17 |  |
| Never heard of physical activity | 16 | 3 | 1 | 20 | 80.0 | 15.0 | 5.0 | 20 |  |
| Total |  |  |  |  |  |  |  |  |  |
| All | 77 | 57 | 7 | 141 | 54.6 | 40.4 | 5.0 |  |  |

Notes: Figures in the parentheses are column relative frequency
Others for place of residence include districts of Rukungiri, Ntungamo, Kiruhura, Kitagwenda, Kabale, Kasese, Rwampara, Kamwenge, Ibanda
Others for occupation include: teacher, health worker, office workers, security guards
The p-values were reported from the Fischer's exact test.

**Table 4:** Significant associated factors to physical activity on bivariate and multivariate adjustment

| Characteristic | Bivariate analysis |  | Multivariate analysis |  |
| --- | --- | --- | --- | --- |
|  | crude OR (cOR),<br>(95% CI) | p-value | adjusted OR (aOR),<br>(95% CI) | p-value |
| **Place of residence** |  |  |  |  |
| Isingiro |  |  |  |  |
| Greater Bushenyi | 0.964(0.751-1.238) | 0.773 | - | - |
| Mbarara |  |  |  |  |
| Others |  |  |  |  |
| **Level of education** |  |  |  |  |
| No formal education |  |  |  |  |
| Completed Primary | 1.420(1.106-1.824) | 0.006 | 1.374(1.055-1.790) | 0.018 |
| Completed Secondary |  |  |  |  |
| Completed Tertiary |  |  |  |  |
| **Heard of physical activity** |  |  |  |  |
| No | 3.934(1.243-12.452) | 0.020 | 3.262(0.953-11.166) | 0.060 |
| Yes |  |  |  |  |
| **Sedentary behaviour** |  |  |  |  |
| More than 6 hours | 0.262(0.123-0.557) | 0.001 | 0.276(0.126-0.606) | 0.001 |
| **Barrier** |  |  |  |  |
| Social influence | 0.970(0.851-1.106) | 0.651 | - | - |

Notes: Others for place of residence include districtis of Rukungiri, Ntungamo, Kiruhura, Kitagwenda, Kabale, Kasese,Rwampara,Kamwenge,Ibanda

## Discussion

This study sought to establish the extent of physical activity and associated factors among patients attending hypertension clinics in Mbarara City, Southwestern Uganda. Physical activity is an effective way of managing hypertension. A participant is physically active if they achieve ≥600 METs in a period of 5 days or more per week (22-23). Therefore, an individual who had moderate or high levels of physical activity was considered to be physically active.

This study found that only 45.39% of the participants were physically active. These findings are in agreement with (24) a study among hypertensive patients attending hospitals in the Central Gander Zone in Addis Ababa who found that (45%) of the participants were physically active. These findings suggest that the majority of hypertensives are physically inactive. Therefore, interventions to increase physical activity among hypertensives should be emphasized. Few participants were active for transport compared to other domains. Creation of low traffic neighborhoods can improve walking and cycling as forms of active transport (25). Therefore, policy formulation to include pedestrian and bicycle lanes during road infrastructure planning may be the solution to solve this transportation physical activity challenge.

Findings of this study reveal that participants who had higher levels of monthly income had high physically activity levels. (26) also found that participants with higher income levels were more physically active. Poverty is more likely to be linked with poor health outcomes, even on the aspect of engaging in physical activity. Therefore, poverty eradication strategies and campaigns can be equally used as platforms for physical activity awareness and counseling.

This study found that those who had information about physical activity were more physically active and contrary to many studies, the study found that those who heard of physical activity from family and friends were more physically active compared to those who heard from health workers therefore health workers may need to assess available family support so as to achieve and sustain physical activity.

36.17% of participants were sedentary (spent more than 6 hours of daily sitting). Almost half of the participants who were physically inactive were also sedentary, (27) reported similar findings. This is a double disaster for people with hypertension (28). Therefore, interventions to improve levels of physical activity should be put in place so as to address this issue.

The current study found level of education was associated with physical activity. Participants who had higher levels of education were more physically active. These results agree with a study by (29) which reported that participants with hypertension who had completed high school and higher levels of education, were more physically active. It has been noted that more educated participants are more aware of the normal values of blood pressure and their significance hence they partake in physical activity. It was also found that, with high education levels, one can easily obtain information, practice counselling, organise and carry out physical activities as per their schedule (24,30). Higher levels of education have also been associated with better economic outcomes (31), which consequently influences physical activity positively (32). Contrasting results where participants with low levels of education were more physically active, have been reported. This was explained in relation to the kind of jobs those with low levels of education engage in that is; more strenuous or physically demanding jobs (17,33).

Approximately, half of the physically inactive participants were also sedentary in the current study. A decrease in physical activity alongside an increase in sedentary behaviour has been reported in other studies (34,27,35). A study among hypertensive subjects reported that each one hour of sedentary behaviour was associated with 0.06mmHg increase in systolic blood pressure and 0.20mmHg or diastolic blood pressure (9) . (36) further noted that higher periods of sedentary behaviour during television viewing are associated with higher risk of death among hypertensive individuals. Therefore, being inactive and sedentary is a double disaster for ill health especially among people with chronic diseases such as hypertension, hence interventions to improve levels of physical activity should be put in place (27).

### Study limitations

This study used subjective study tools therefore the results were subject to recall bias. The study tool adopted only assessed physical activity of individuals aged 18-69 years only therefore our results cannot be used to inform possible interventions and recommendations for improving physical activity among patients younger than 18 years and older than 69 years of age. The study sample size was small which makes our results unrepresentative to the general population.

### Conclusion and Recommendations

Our study revealed that majority of hypertensive patients was physically inactive despite physical activity benefits. Almost half of the hypertensive patients were both physically inactive and sedentary, each of which has an independent negative effect on blood pressure control. Therefore, programs or interventions informing of the benefits of physical activity in management of hypertension should be greatly emphasized in the local settings. We recommend that, urban planners and ministry of transport provide for pedestrian and cyclists’ lanes during road construction, and for ministry of health to establish cardiac rehabilitation programs in all government hospitals for all patients with hypertension and other cardiovascular diseases.

## Data Availability

the data refereed to in the manuscript is available whenever needed for use

## Acknowledgements

We want to acknowledge all those who have made this research project successful. We first thank God. Secondly, we thank our parents who have offered the necessary support. We also want to acknowledge our supervisor Dr. Arubaku Wilfred who guided us throughout the project. Many thanks to the MUST Research Ethics Committee for granting approval of our proposal, DMA Diagnostics and Laboratory Limited and Mbarara Medical Specialists Clinic for granting us opportunities to undertake research at their clinics. We also acknowledge Ms Eleanor Turyakira and Dr. Muwanguzi Moses for the support rendered to us during the data analysis process. Further thanks to the participants who participated in this study.

